# Comparative Evaluation of a System One Model and a General-Purpose Large Language Model on the Korean Physical Therapist Licensing Examination

**DOI:** 10.64898/2026.09.26.26364067

**Authors:** Jun-hee Kim

## Abstract

**Background:** System One Model is designed for structured decision-making and can provide probabilistic outputs, but their performance in domain-specific physical therapy tasks has not been established.

**Objective:** To evaluate the performance and potential utility of Jev, a System One Model, for professional knowledge-based selection tasks in physical therapy by comparing it with GPT-4o.

**Methods:** A total of 380 multiple-choice questions from the 2024 and 2025 Korean Physical Therapist Licensing Examinations were independently submitted to Jev and GPT-4o. The outcomes included answer accuracy, subgroup performance, output validity, API response latency, token usage, estimated API cost, and Jev’s prediction uncertainty. A retrospective probability-based model-cascading simulation was also performed.

**Results:** Jev correctly answered 282 questions (74.2%), whereas GPT-4o answered 326 (85.8%), a difference of -11.6 percentage points. Jev produced valid structured responses for all questions, whereas GPT-4o produced three invalid responses. Jev’s selected-answer probabilities discriminated correct from incorrect answers (AUROC = 0.866), and accuracy reached 99.3% when probabilities were greater than or equal to 0.90. The median API response latency was 0.94 s for Jev and 1.06 s for GPT-4o, with estimated costs of US$0.008 and US$0.161, respectively. At a retrospective cascading threshold of 0.70, accuracy was 86.6% with 175 simulated GPT-4o calls.

**Conclusion:** Jev showed lower overall accuracy than GPT-4o but provided reliable structured outputs, informative prediction probabilities, lower latency, and lower estimated cost. Its probabilistic outputs warrant further prospective evaluation for selective model escalation and decision-support workflows in physical therapy.

## INTRODUCTION

Clinical decision-making in physical therapy is a complex process that requires physical therapists to integrate patients’ medical histories, symptoms, and physical examination findings to determine appropriate assessment and intervention strategies [1–3]. Clinical reasoning involves the integration of professional knowledge and individual patient characteristics, and is continuously adapted and refined in response to changes in patients’ conditions and treatment outcomes [2]. With recent advances in artificial intelligence (AI), various AI-based technologies have been investigated for use in physical rehabilitation to support patient assessment, treatment, and rehabilitation processes [4–6]. In particular, AI has the potential to assist physical therapists in interpreting patient-related information and making informed clinical decisions [5,7]. However, evidence regarding the clinical effectiveness of AI-based rehabilitation technologies remains limited, highlighting the need for systematic evaluation before their integration into clinical practice [4,8].

Recent advances in large language models (LLMs) have increased interest in their potential to support professional knowledge-based decision-making in healthcare and rehabilitation [9,10]. However, general-purpose LLMs typically generate responses by predicting tokens sequentially, and their use in tasks requiring selection from predefined alternatives may require additional instructions or output constraints to ensure that the responses follow a specified format[11–13]. Such constraints may affect task performance, and response latency and computational costs are also important considerations when evaluating these models for practical use [11,14,15]. Furthermore, the reliability of the predicted probabilities generated by LLMs in medical applications warrants evaluation alongside the correctness of their predictions [16]. Together, these considerations motivate the investigation of models designed to return structured outputs and associated predicted probabilities, rather than unrestricted textual responses.

TypeSafe AI recently launched Jev, its first publicly available System One Model, which is designed to perform structured decision-making tasks rather than generate unrestricted textual responses [17]. Jev processes information describing a given situation (state) and performs predefined tasks through three task types: choice, score, and binary judgment (noul). Unlike the sequential text generation approach used by general-purpose LLMs, Jev produces type-safe structured outputs with predicted probabilities and confidence scores. According to TypeSafe AI, Jev is trained using Reinforcement Learning for Calibrated Decisions (RLCD), a training approach intended to optimize the calibration of model decisions rather than human preference for generated responses. Consequently, Jev is designed to explicitly represent uncertainty through probabilistic outputs, enabling downstream systems to use confidence thresholds when determining whether to accept, review, or escalate model decisions. Through its choice function, the model selects an option from a predefined set and returns a predicted probability distribution of the available options. These characteristics may facilitate its integration into structured decision-making workflows, including physical therapy tasks that require the selection of appropriate assessment or intervention strategies.

Although Jev provides structured outputs and predicted probabilities, these technical features alone do not establish its ability to make accurate and efficient decisions in domain-specific tasks. Because general-purpose LLMs can also be instructed to select among predefined alternatives, a direct comparison is necessary to characterize the performance of Jev under equivalent decision-making tasks [12]. In physical therapy, such tasks encompass diverse forms of professional knowledge and judgment, ranging from interpreting assessment findings to selecting and applying appropriate interventions [2]. Therefore, overall accuracy alone may not adequately reflect differences in answer accuracy across different types of physical therapy decision-making tasks. Furthermore, previous research has demonstrated that the predicted probabilities generated by AI models do not necessarily correspond to their observed accuracy, highlighting the importance of evaluating and communicating prediction uncertainty in healthcare applications [18,19]. Consequently, exploring the potential and limitations of Jev in physical therapy requires a multidimensional evaluation of its answer accuracy overall and across different question types, API response latency, and output validity, together with an assessment of the relationship between its predicted probabilities and answer correctness.

Accordingly, this study aimed to explore the potential and limitations of Jev, a System One Model, for professional knowledge-based selection tasks in physical therapy through a direct comparison with GPT-4o, a general-purpose large language model (LLM). Using questions from the 2024 and 2025 Korean Physical Therapist Licensing Examinations as standardized evaluation tasks, the models’ answer accuracy overall and across different question types, including questions with patient-specific information, were examined. API response latency, output validity, token usage, and estimated API costs were also evaluated to characterize the practical considerations associated with their use in structured selection tasks. In addition, the relationship between the probabilities assigned to Jev’s selected answers and their answer correctness was explored, along with the potential utility of probability-based model cascading through a retrospective simulation. This evaluation was intended to identify the performance characteristics and limitations of Jev as a System One Model and provide a basis for further investigation of structured AI decision-making approaches in physical therapy.

## METHODS

### 1. Study Design

This study was designed to compare the performance of Jev, a System One Model, and GPT-4o, a general-purpose large language model, on professional knowledge-based selection tasks in physical therapy. A total of 380 multiple-choice questions from the 2024 and 2025 Korean Physical Therapist Licensing Examinations were used as the standardized evaluation tasks. Each question was independently presented to both models under predefined experimental conditions, and their responses were compared using the official final answer key. To examine answer accuracy across different question types, the examination questions were classified according to their primary knowledge and decision-making requirements, with additional consideration of whether patient-specific information was provided.

**Figure 1.**
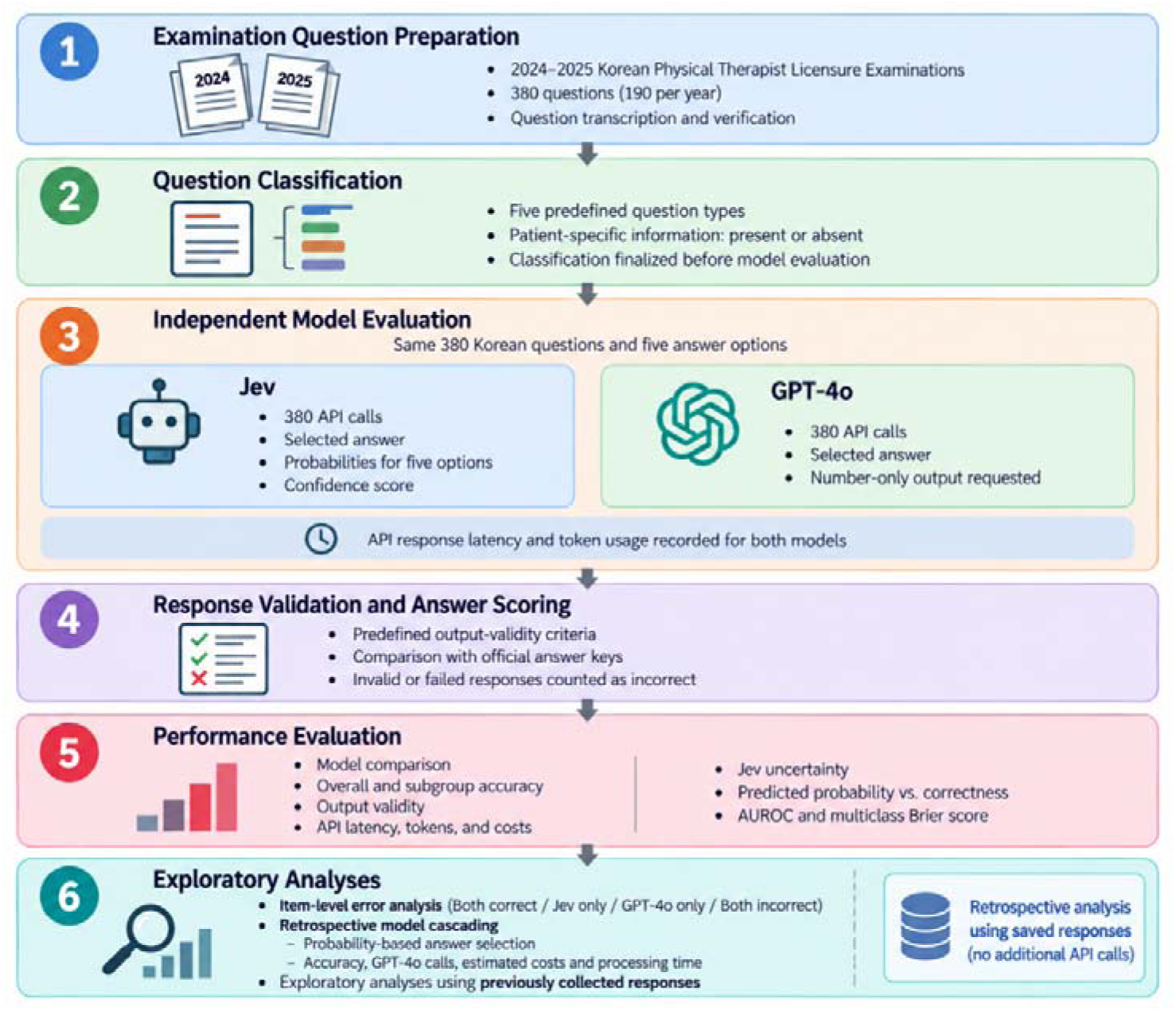
Study Workflow

### 2. Examination Questions and Classification

A total of 380 publicly available written examination questions from the 2024 and 2025 Korean Physical Therapist Licensing Examinations were included in this study. For each examination year, 190 written questions were publicly released by the Korea Health Personnel Licensing Examination Institute (KHPLEI), comprising four subjects: Physical Therapy Fundamentals (60 questions), Physical Therapy Diagnosis and Assessment (45 questions), Physical Therapy Intervention (65 questions), and Medical Laws and Regulations (20 questions). The 70 practical examination questions administered each year were not publicly released by the KHPLEI and therefore could not be included in the analysis. All included questions followed a five-option, single-best-answer format. The original Korean wording was preserved, and the official final answer keys were used as the reference standard. All 380 publicly available questions could be presented in text format without requiring additional visual information. To evaluate performance across different types of professional knowledge-based selection tasks, the questions were classified into five question types according to their primary knowledge and decision-making requirements: (1) Fundamental and Professional Knowledge, (2) Condition and Assessment Judgment, (3) Intervention and Action Selection, (4) Intervention Application and Adjustment, and (5) Laws and Regulations. Each question was assigned to a single question type based on its primary task. Questions were additionally classified according to whether they contained patient-specific information, including patient characteristics, medical history, symptoms, or examination findings. Question classification was performed according to the predefined criteria by the investigator, who had more than 10 years of professional experience in physical therapy.

### 3. AI Models and Experimental Configuration

Two models were evaluated in this study: Jev (jev-1.13.0; TypeSafe AI), a System One Model, and GPT-4o (gpt-4o-2024-08-06; OpenAI), a general-purpose large language model. Jev was evaluated using its choice function, which selects an answer from predefined answer options and returns a predicted probability distribution across the five answer options. GPT-4o was selected as the reference model to compare the performance of a general-purpose LLM with that of a System One Model. Both models were accessed through their respective API. Each model received the same Korean question stem and five answer options, with instructions to select the single most appropriate answer. For Jev, the question was provided as the state, and five answer options were defined as choice criteria. For GPT-4o, the question and answer options were provided through the Chat Completions API with instructions to return only the number of the selected answer without additional explanation. GPT-4o was configured with a temperature of 0, random seed of 42, and maximum output length of five tokens. Each question was submitted independently to both models, without additional prompts or follow-up interactions.

### 4. Experimental Procedure

All experiments were conducted on September 23, 2026, using a single computer and network environment. Both models were accessed through their respective APIs using a Python-based experimental pipeline. The 380 examination questions were processed sequentially, with each question submitted once to each model. To minimize the potential effects associated with the order of API requests, the API call order was alternated across consecutive questions. Jev was called first for one question, followed by GPT-4o, whereas GPT-4o was called first for the subsequent question. This procedure ensured that each model was called first for an equal number of questions. All 760 API calls were completed within a single experimental session without parallel processing or additional follow-up interactions. Each API call was assigned a timeout of 60 seconds. A predefined retry procedure was implemented for transient API errors, including rate-limit errors, server errors, and network failures, allowing up to five attempts with exponential backoff. Responses were recorded immediately after each API call to preserve the original results and prevent duplicate requests. For each question, the selected answer, response status, API response latency, model version, and token usage were recorded. In addition, the predicted probability distribution across the five answer options and the confidence score were collected from Jev. Model responses were processed using predefined parsing rules to extract the selected answer and determine whether the response satisfied the required output format. Invalid responses were recorded without manual correction or additional model queries.

### 5. Performance Evaluation

The primary outcome was answer accuracy, defined as the proportion of correctly answered questions based on official final answer keys. The overall accuracy was calculated for each model across all 380 questions, with invalid responses and failed API calls counted as incorrect. Answer accuracy was further evaluated based on the examination year, examination subject, question type, and presence of patient-specific information. Answer accuracy and differences in accuracy were reported with 95% confidence intervals (CIs) estimated from 2,000 item-level bootstrap resamples, preserving paired responses to each question. Output validity was assessed by determining whether Jev returned a selected answer with predicted probabilities for all five answer options and whether GPT-4o returned a response containing exactly one answer between 1 and 5. Answer accuracy was calculated for valid responses. Prediction uncertainty was evaluated for Jev by examining the relationship between the selected-answer probability, defined as the probability assigned to Jev’s selected answer, and answer correctness. Probability calibration was assessed descriptively by dividing the selected-answer probabilities into five predefined probability intervals (below 0.40, 0.40 to less than 0.60, 0.60 to less than 0.80, 0.80 to less than 0.90, and 0.90 to 1.00) and comparing the mean predicted probability with the observed accuracy in each interval. Discrimination between correct and incorrect answers was evaluated using the area under the receiver operating characteristic curve (AUROC), with the selected-answer probability as the predictor. The multiclass Brier score was used to evaluate the overall quality of the predicted probability distributions across all five answers. Jev’s confidence scores were analyzed separately from the selected-answer probabilities. The API operational performance was evaluated using API response latency, token usage, and estimated API costs. API response latency was measured from API request initiation to receipt of the complete response, including network transmission and server-processing time. The estimated API costs were calculated using each provider’s API list prices. These measures reflect the operational characteristics of API services rather than the intrinsic computational efficiency of the models.

### 6. Error Analysis and Model Cascading

Item-level correctness patterns were analyzed by classifying questions into four groups according to the correctness of the two models’ responses: both models correct, Jev only correct, GPT-4o only correct, and both models incorrect. The distribution of these groups was examined to characterize the overlap and differences in model errors. The potential utility of probability-based model cascading was evaluated through a retrospective model-cascading simulation using previously collected responses. Jev’s answer was retained when its selected-answer probability met or exceeded a specified threshold (hereafter, the selected-answer probability threshold); otherwise, GPT-4o’s answer was used. Six probability thresholds (0.50, 0.60, 0.70, 0.80, 0.90, and 0.95) were examined. For each threshold, answer accuracy, the number of simulated GPT-4o API calls, and estimated API costs were calculated. The estimated total processing time was calculated by summing the observed API response latencies of the API calls required under each threshold. Because the thresholds were selected after the primary results had been examined, this analysis was considered exploratory.

## RESULTS

### 1. Answer Accuracy

Across the 380 examination questions, Jev correctly answered 282 questions (74.2%; 95% CI: 69.7–78.4%), whereas GPT-4o correctly answered 326 questions (85.8%; 95% CI: 82.4–89.2%) (Table 1). The difference in accuracy between Jev and GPT-4o was −11.6 percentage points (95% CI: −16.3 to −7.1 percentage points). In the 2024 examination, answer accuracy was 72.1% for Jev and 84.2% for GPT-4o; the corresponding values for the 2025 examination were 76.3% and 87.4%, respectively. GPT-4o also showed higher accuracy across all four examination subjects. Subject-specific accuracy ranged from 65.6% to 78.5% for Jev and from 77.5% to 90.0% for GPT-4o.

**Table 1.** Overall and Subgroup Answer Accuracy.

| Group | n | Jev, n (%)<br>[95% CI] | GPT-4o, n<br>(%) [95%<br>CI] | Difference,<br>pp (95% CI) | Correct only<br>for Jev /<br>GPT-4o |
| --- | --- | --- | --- | --- | --- |
| All questions | 380 | 282 (74.2)<br>[69.7–78.4] | 326 (85.8)<br>[82.4–89.2] | –11.6 (–16.3<br>to –7.1) | 18 / 62 |
| 2024 examination | 190 | 137 (72.1)<br>[65.3–78.4] | 160 (84.2)<br>[78.9–88.9] | –12.1 (–18.9<br>to –5.8) | 12 / 35 |
| 2025 examination | 190 | 145 (76.3)<br>[70.5–82.1] | 166 (87.4)<br>[82.6–92.1] | –11.1 (–16.8<br>to –5.3) | 6 / 27 |
| Physical Therapy<br>Fundamentals | 120 | 94 (78.3)<br>[70.8–85.0] | 108 (90.0)<br>[84.2–95.0] | –11.7 (–19.2<br>to –4.2) | 4 / 18 |
| Physical Therapy<br>Diagnosis and<br>Assessment | 90 | 59 (65.6)<br>[55.6–75.6] | 73 (81.1)<br>[72.2–88.9] | –15.6 (–26.7<br>to –4.4) | 7 / 21 |
| Physical Therapy<br>Intervention | 130 | 102 (78.5)<br>[70.8–85.4] | 114 (87.7)<br>[81.5–93.1] | –9.2 (–16.9<br>to –2.3) | 7 / 19 |
| Medical Laws and<br>Regulations | 40 | 27 (67.5)<br>[52.5–80.0] | 31 (77.5)<br>[65.0–90.0] | –10.0 (–20.0<br>to –2.5) | 0 / 4 |

**Note.** Invalid responses and failed API calls were considered incorrect (Jev: 0; GPT-4o: 3). The 95% CIs were calculated using 2,000 item-level bootstrap resamples with paired bootstrap resamples for differences in accuracy. Differences are expressed as Jev minus GPT-4o in percentage. CI, confidence interval; pp, percentage points.

### 2. Answer Accuracy by Question Type

GPT-4o showed higher answer accuracy than Jev across all five question types (Table 2). The differences in accuracy ranged from 9.6 to 13.1 percentage points. The largest observed difference was in Fundamental and Professional Knowledge (Jev: 76.5%; GPT-4o: 89.6%), whereas the smallest was in Intervention and Action Selection (Jev: 73.1%; GPT-4o: 82.7%). The Intervention Application and Adjustment question type contained only 19 questions, resulting in a relatively wide confidence interval for the accuracy difference. Among the 61 questions with patient-specific information, Jev and GPT-4o achieved accuracies of 82.0% and 83.6%, respectively, with a difference of −1.6 percentage points (95% CI: −14.8 to 11.5 percentage points). Among the 319 questions without patient-specific information, the corresponding accuracies were 72.7% and 86.2%, with a difference of −13.5 percentage points (95% CI: −18.2 to −8.8 percentage points).

**Table 2.** Answer Accuracy by Question Type and Patient-Specific Information.

| Group | n | Jev, n (%)<br>[95% CI] | GPT-4o, n<br>(%) [95% CI] | Difference,<br>pp (95% CI) | Correct only<br>for Jev /<br>GPT-4o |
| --- | --- | --- | --- | --- | --- |
| Fundamental and<br>Professional<br>Knowledge | 183 | 140 (76.5)<br>[70.5–82.5] | 164 (89.6)<br>[85.2–94.0] | –13.1 (–19.7<br>to –6.6) | 8 / 32 |
| Condition and<br>Assessment Judgment | 86 | 64 (74.4)<br>[65.1–83.7] | 73 (84.9)<br>[76.7–91.9] | –10.5 (–20.9<br>to 0.0) | 6 / 15 |
| Intervention and<br>Action Selection | 52 | 38 (73.1)<br>[61.5–84.6] | 43 (82.7)<br>[73.1–92.3] | –9.6 (–21.2 to<br>+1.9) | 3 / 8 |
| Intervention<br>Application and<br>Adjustment<sup>a</sup> | 19 | 13 (68.4)<br>[47.4–89.5] | 15 (78.9)<br>[57.9–94.7] | –10.5 (–31.6<br>to +10.5) | 1 / 3 |
| Laws and Regulations | 40 | 27 (67.5)<br>[52.5–80.0] | 31 (77.5)<br>[65.0–90.0] | –10.0 (–20.0<br>to –2.5) | 0 / 4 |
| Without patient-specific information | 319 | 232 (72.7)<br>[67.7–77.4] | 275 (86.2)<br>[82.4–90.0] | –13.5 (–18.2<br>to –8.8) | 11 / 54 |
| With patient-specific<br>information | 61 | 50 (82.0)<br>[70.5–91.8] | 51 (83.6)<br>[73.8–91.8] | –1.6 (–14.8 to<br>+11.5) | 7 / 8 |

**Note.** Invalid responses and failed API calls were considered incorrect. The 95% CIs were calculated using 2,000 item-level bootstrap resamples to preserve paired responses. Differences are expressed as Jev minus GPT-4o in percentage. Results for Intervention Application and Adjustment should be interpreted with caution due to the small sample size (n = 19). CI, confidence interval; pp, percentage points.

### 3. Output Validity and Prediction Uncertainty

Jev returned valid responses for all 380 questions, whereas GPT-4o produced three invalid responses (0.8%) under the predefined output criteria. All three invalid responses contained an answer number followed by additional numerical information. When only valid responses were considered, the answer accuracy was 74.2% for Jev and 86.5% for GPT-4o. For Jev, the median selected-answer probability was 0.905 for correctly answered questions and 0.460 for incorrectly answered questions. The observed accuracy generally increased across the predefined probability intervals, reaching 99.3% among the 146 questions with selected-answer probabilities of at least 0.90 (Figure 2). The AUROC of the selected-answer probabilities for distinguishing correct from incorrect answers was 0.866, and the multiclass Brier score was 0.342. The AUROC of Jev’s confidence scores was also 0.866.

**Figure 2.**
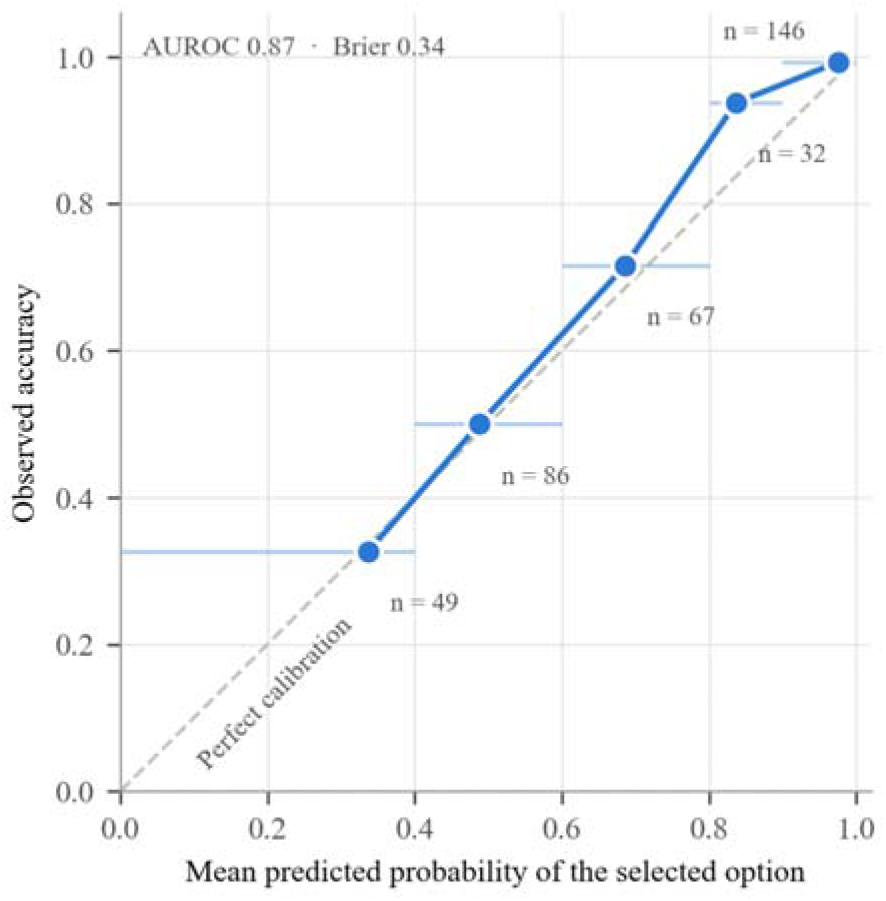
Calibration of Jev’s Selected-Answer Probabilities

### 4. API Operational Performance

Jev had a median API response latency of 0.94 s (interquartile range [IQR]: 0.89–1.02 s), compared with 1.06 s (IQR: 1.00–1.25 s) for GPT-4o (Table 3). Across the 380 questions, Jev used 182,102 input tokens and 19,760 output tokens, whereas GPT-4o used 62,612 input tokens and 423 output tokens. Despite its higher token usage, Jev had a lower estimated API cost (US$0.008) than GPT-4o (US$0.161). These values represent the observed API response latency and estimated API costs under the experimental conditions.

**Table 3.** API Operational Performance of Jev and GPT-4o.

| Metric | Jev | GPT-4o |
| --- | --- | --- |
| API response latency, median [Q1, Q3], s | 0.94 [0.89, 1.02] | 1.06 [1.00, 1.25] |
| API response latency, mean / maximum, s | 1.01 / 2.17 | 1.20 / 10.79 |
| Input tokens, total (mean per question) | 182,102 (479.2) | 62,612 (164.8) |
| Output tokens, total | 19,760 | 423 |
| Estimated API cost for 380 questions, USD | 0.008 | 0.161 |
| Invalid responses, n (%) | 0 (0.0%) | 3 (0.8%) |
| Failed API calls / questions requiring retries, n | 0 / 0 | 0 / 0 |
| Accuracy among valid responses, % (n) | 74.2 (380) | 86.5 (377) |
| Model version | jev-1.13.0 | gpt-4o-2024-08-06 |

**Note.** API response latency was measured end-to-end by the client (request sent to full response received) and included network and server queuing; it was not the inference time of the model. All 760 API calls were made sequentially from a single process within a 14-minute window with an alternating API call order. The estimated API costs were calculated using API list prices at the time of the study (Jev: US\$0.042 per million input tokens, output not charged; GPT-4o: US\$2.50 per million input tokens and US\$10.00 per million output tokens). Q1, first quartile; Q3, third quartile.

### 5. Error Analysis and Model Cascading

Both models answered 264 of the 380 questions correctly, whereas both answered 36 questions incorrectly (Figure 3). Jev alone answered 18 questions correctly, compared with 62 questions answered correctly by GPT-4o alone. In the retrospective model-cascading simulation, the answer accuracy ranged from 81.8% to 86.6% across the six selected-answer probability thresholds (Figure 4). At a selected-answer probability threshold of 0.70, the retrospective model-cascading simulation achieved an accuracy of 86.6%, with 175 simulated GPT-4o API calls and an estimated API cost of US$0.085. At a selected-answer probability threshold of 0.80, accuracy was 85.8%, with 202 simulated GPT-4o API calls and an estimated API cost of US$0.096. The estimated total processing time was longer than that of GPT-4o alone across all evaluated thresholds.

**Figure 3.**
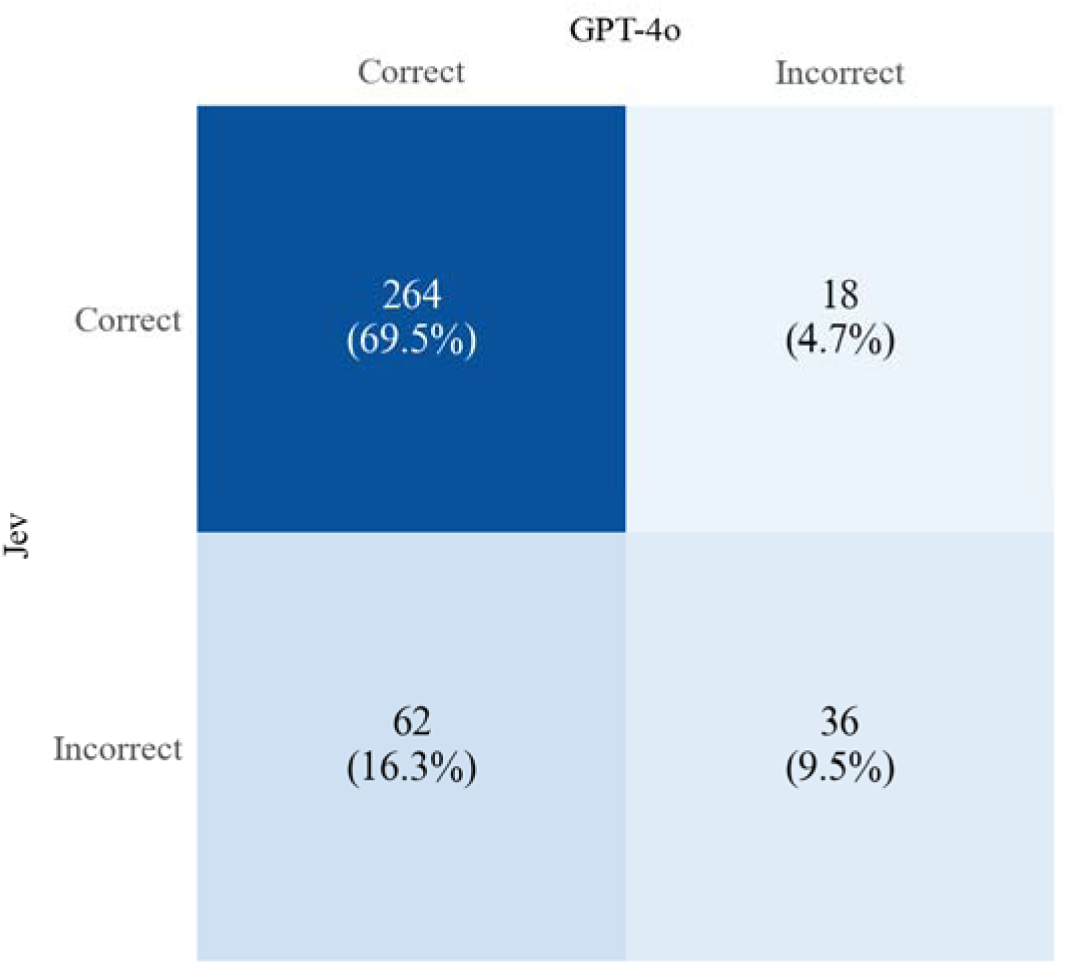
Item-Level Correctness Patterns

**Figure 4.**
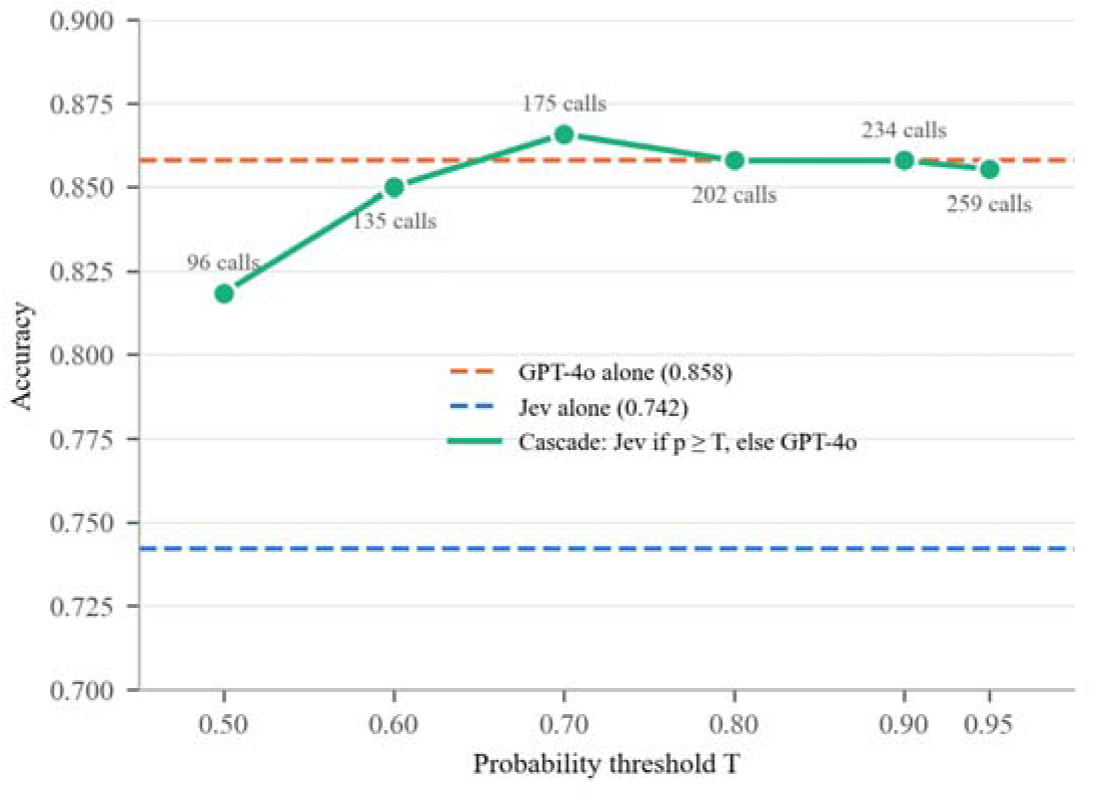
Performance of Retrospective Model Cascading

## DISCUSSION

This study evaluated the performance and potential utility of Jev, a System One Model, in professional knowledge-based selection tasks in physical therapy through a direct comparison with GPT-4o, a general-purpose large language model. Jev achieved an overall accuracy of 74.2%, compared with 85.8% for GPT-4o, representing a difference of 11.6 percentage points. GPT-4o demonstrated higher accuracy across all question types, although the difference between the two models was smaller for questions with patient-specific information than for other questions. Jev returned valid responses for all questions, and its selected-answer probabilities provided useful information for distinguishing correct answers from incorrect answers. Jev also showed lower API response latency and estimated API costs than GPT-4o did. These findings indicate that although Jev demonstrated lower accuracy than GPT-4o in professional knowledge-based selection tasks, it exhibited distinct characteristics in terms of structured outputs, predicted probabilities, and API operational performance.

The accuracy achieved by GPT-4o in the present study was consistent with previous findings from healthcare Licensing examination. Kim and Shin (2025) reported an accuracy of 88.9% for GPT-4o on 960 Korean Physical Therapist Licensing Examination questions [20]. Similarly, Woo et al. (2026) evaluated ChatGPT-4o on 193 questions from the 59th Japanese National Examination for Physical Therapists and reported an accuracy of 80.0%. Sawamura et al. (2024) evaluated ChatGPT 4.0 across 980 questions from five Japanese National Examinations for Physical Therapists and reported an overall accuracy of 73.4%, which increased to 80.5% for text-only questions [21,22]. Kim et al. (2025) also reported 83.2% accuracy for GPT-4o on 942 Korean medical Licensing examination questions [23]. These findings demonstrate that general-purpose LLMs can achieve high accuracy in professional knowledge-based selection tasks across various healthcare disciplines. In contrast, Jev demonstrated lower accuracy than GPT-4o in the present study, despite being designed for structured selection tasks, suggesting that structured output capabilities do not necessarily ensure high accuracy in domain-specific knowledge-based decision-making. However, because the differences in the training data, model architecture, and internal processing mechanisms were not controlled, the observed performance difference cannot be attributed to any specific factor.

The observed difference in accuracy between Jev and GPT-4o also varied descriptively according to question characteristics. GPT-4o demonstrated higher accuracy than Jev across all five predefined question types. Among questions containing patient-specific information, the observed difference between the models was numerically smaller than that among questions without patient-specific information. However, this subgroup included only 61 questions, and the confidence interval for the accuracy difference was wide. Therefore, the present findings do not establish that the relative performance of the two models differs according to the presence of patient-specific information. Huhn et al. (2019) described clinical reasoning in physical therapy as a process that integrates professional knowledge, patient characteristics, and clinical context [2]. Similarly, Hao et al. (2025) evaluated ChatGPT-4 across the subjective, objective, assessment, and planning stages of 10 musculoskeletal physical therapy cases and found that performance varied across stages, with the lowest accuracy observed during the objective examination component [24]. These findings indicate that model performance may vary according to the information provided and the type of judgment required. However, because the question type, examination subject, and presence of patient-specific information may overlap, the observed subgroup patterns should be interpreted descriptively. Furthermore, performance on Licensing examination questions does not directly reflect real-world clinical decision-making ability.

A key characteristic of Jev is its ability to select an answer from predefined answer options while providing a predicted probability distribution across those options. In the present study, Jev’s selected-answer probabilities demonstrated the ability to distinguish correct from incorrect answers (AUROC = 0.866), with an accuracy of 99.3% for questions with selected-answer probabilities of at least 0.90. Although predicted probabilities do not necessarily correspond to observed accuracy (Guo et al., 2017), such information may support the identification of questions requiring additional review [18]. Kompa et al. (2021) highlighted the importance of uncertainty quantification in medical AI and discussed the potential for models to abstain from uncertain predictions and refer such cases for additional expert review [19]. Similarly, Lee et al. (2025) demonstrated that a confidence-linked, uncertainty-based staged LLM framework reduced the need for manual chart review by approximately 75% while maintaining high overall performance through selective expert review of uncertain cases [25]. These findings suggest that Jev’s structured outputs and selected-answer probabilities could be incorporated into clinical decision-support workflows to identify cases requiring additional professional review. In physical therapy, such a framework may support the preliminary selection or prioritization of predefined assessment or intervention options, while final clinical decisions remain subject to independent evaluation by physical therapists. Furthermore, the retrospective model-cascading simulation suggested that Jev’s selected-answer probabilities could be used to determine when a GPT-4o response would be requested. At selected probability thresholds, this approach reduced the number of simulated GPT-4o API calls and estimated API costs while producing accuracy point estimates similar to those of GPT-4o alone. However, because the cascading strategy was evaluated retrospectively using the same set of previously collected responses and the thresholds were explored after examination of the primary results, these findings should be considered exploratory rather than evidence of equivalent or superior performance. Independent validation using prospectively defined thresholds and sequential API execution is required before the operational effectiveness of this approach can be established.

This study had several limitations that should be considered. First, the evaluation was based exclusively on text-only questions from the Korean Physical Therapist Licensing Examinations, with all questions presented in the original Korean. Therefore, the findings may not be generalizable to examinations or decision-making tasks conducted in other languages, particularly because Jev’s language-specific performance has not yet been extensively established. In addition, Licensing examination questions may not fully represent the complexity of real-world clinical decision-making, which involves diverse patient information and dynamic clinical conditions. Second, only two specific model versions were evaluated, and each question was primarily assessed using a single API call within one experimental session. Therefore, the generalizability of the findings across different models, repeated evaluations and operational environments is uncertain. In addition, the comparison of output validity should be interpreted in the context of the different output mechanisms used for the two models, as Jev returned structured choice outputs, whereas GPT-4o generated text responses under the instruction to return only a single answer number. Additionally, the potential overlap between publicly available examination questions and the models’ training data could not be verified. Third, the relatively small number of questions in certain subgroups limited the interpretation of differences in accuracy across question types and patient-specific information. Finally, the probability-based model-cascading strategy was evaluated retrospectively using previously collected responses, and its performance was validated using independent datasets and prospective sequential model execution. Future studies should evaluate Jev across multiple languages and use clinically representative tasks, including patient assessment, intervention selection, and treatment adjustment, to further investigate its potential utility in physical therapy clinical decision support systems.

## CONCLUSION

This study compared Jev, a System One Model, with GPT-4o on professional knowledge-based selection tasks using Korean Physical Therapist Licensing Examination questions. Jev demonstrated lower overall answer accuracy than GPT-4o but provided consistently valid structured outputs and selected-answer probabilities that effectively distinguished correct from incorrect responses. Its lower API response latency and estimated API cost, together with the exploratory retrospective model-cascading results, suggest that its probabilistic outputs may be useful for further investigation in selective model-escalation workflows. However, the cascading findings were derived from a retrospective simulation and should not be interpreted as evidence of equivalent or superior performance to GPT-4o alone. Further studies using independent datasets, prospectively defined thresholds, prospective cascading designs, and clinically representative physical therapy tasks are needed to determine the practical value of System One Models in decision-support settings.

## Data Availability

The examination questions and official answer keys used in this study are publicly available from the Korea Health Personnel Licensing Examination Institute (KHPLEI). The model-generated responses and data supporting the findings of this study are available from the corresponding author upon reasonable request.

https://www.kuksiwon.or.kr/CollectOfQuestions/brd/m_116/list.do

